# Acceptability of community-based maternal and newborn care in South Sudan: A qualitative study using the Theoretical Framework of Acceptability

**DOI:** 10.64898/2026.05.14.26353170

**Authors:** Lual Agok Luka, Teresia Macharia, Grace Kimemia, Geeta Nanda, Alom Atak Ayom, Amijong Deng, James Majok Dut Kuol, Muna Jama, Lual Mayom Nyuany, Inna Caroline, Kadra Noor, Naoko Kozuki

## Abstract

South Sudan faces among the highest maternal and newborn mortality rates globally, with approximately 87% of deliveries occurring at home without skilled birth attendance. In 2024, the International Rescue Committee launched a Community-Based Maternal and Newborn Care (CBMNC) program in Aweil East County, Northern Bahr El Ghazal, deploying trained Boma Health Workers (BHWs) to deliver essential maternal and newborn health services at the household level. This study explored the acceptability of the CBMNC model among diverse stakeholders. This qualitative descriptive study was grounded in the Theoretical Framework of Acceptability (TFA). Data were collected between May and July 2025 through 17 focus group discussions (FGDs), 14 in-depth interviews (IDIs), and 10 key informant interviews (KIIs) with 185 participants, including program recipients, male partners, mothers and mothers-in-law, Boma and Hospital Health Committee (BHC/HHC) members, BHWs, supervisors, and health system stakeholders at state and national levels. Framework analysis combining deductive coding based on the seven TFA constructs with inductive thematic analysis was used. CBMNC was well accepted by recipients and their families, despite provider and health system concerns about sustainability. Trust in community-selected BHWs made home-based care valuable, especially given limited facility access. Intervention coherence relied on pictorial aids, repeated visits, and peer learning to address low literacy. Participants perceived commodity interventions like misoprostol and chlorhexidine as impactful, while behavioral counseling was less recognized. Clients faced minimal burden, but providers experienced significant challenges and inadequate compensation. Health stakeholders were cautiously optimistic but questioned lay provider capacity and long-term viability in a fragile environment. CBMNC can achieve high community acceptability when delivered through trusted, community-selected health workers using contextually appropriate strategies. However, community acceptability alone is insufficient for sustainable scale-up. Addressing provider compensation, workload, and structural integration into national health systems is essential to ensure that gains in acceptability translate into sustained service delivery.

## Introduction

As nations strive to achieve the Sustainable Development Goals (SDGs) and address persistent health equity gaps, evidence from low- and middle-income countries (LMICs) continues to affirm the transformative power of community-led care. Community Health Workers (CHWs), as trusted members residing within their communities, have demonstrated effectiveness in enhancing access to healthcare services, improving health outcomes, and strengthening community trust in health systems. Specifically for maternal and newborn health (MNH), systematic reviews have shown that community-based intervention packages reduced neonatal mortality by 25%, stillbirths by 19%, and perinatal mortality by 22%, while also increasing institutional deliveries and early breastfeeding [1] that community-based educational interventions significantly reduced neonatal and perinatal mortality, improved antenatal care uptake, and increased early breastfeeding initiation [2]. Similarly, CHWs delivering preventive interventions, such as health education, essential newborn care, and psychosocial support, were effective in promoting mother-performed practices like skin-to-skin care and exclusive breastfeeding. CHWs’ unique position as culturally competent intermediaries fosters trust and rapport, enabling them to bridge gaps between health systems and marginalized communities [3].

Despite this strong evidence, community-based MNH programs remain chronically underfunded and deprioritized, particularly in fragile and conflict-affected settings where the need is greatest. Strategic investment in these interventions may be able to enhance progress toward global health targets, reduce preventable deaths, and build resilient systems capable of delivering care to the most underserved populations.

South Sudan, as the world’s newest nation experiencing recurrent conflict, underscores the critical need for targeted investments in community-based MNH services within fragile and conflict-affected settings. The country reports a maternal mortality ratio of 1,223 deaths per 100,000 live births, one of the highest globally [4]. Approximately 87% of deliveries occur at home, often without the presence of skilled birth attendants or access to emergency obstetric care [5]. The consequences for newborns are equally severe, with a neonatal mortality rate of 40 deaths per 1,000 live births and a stillbirth rate of 26 per 1,000 births [6]. Access to life-saving interventions remains critically low, with only 0.6% of expectant mothers having access to comprehensive emergency obstetric and neonatal services [7]. These poor health outcomes reflect the lifelong effects of geographic isolation, economic hardship, cultural barriers, and systemic weaknesses in the health system. In such contexts, community-centered interventions may help bridge some critical gaps to life-saving care.

While the evidence base for community-based MNH interventions has grown considerably, evaluations have predominantly focused on effectiveness and feasibility, with comparatively limited attention to acceptability which is the degree to which services are perceived as appropriate, culturally aligned, and trustworthy by the communities they serve [8]. This gap is consequential because acceptability is a prerequisite for intervention fidelity and sustainable scale-up; programs that are not perceived as acceptable by recipients, providers, and health system actors are unlikely to be adopted consistently or maintained over time, regardless of their demonstrated efficacy [8]. Furthermore, although the Theoretical Framework of Acceptability (TFA) offers a comprehensive, multi-construct approach to assessing acceptability, it has rarely been applied systematically in fragile and conflict-affected settings in sub-Saharan Africa, where contextual factors such as insecurity, displacement, low literacy, and weakened health systems may shape acceptability in distinct ways.

This study addressed these gaps by exploring the acceptability of the CBMNC model in Aweil East County using the TFA, capturing perspectives from women, families, local stakeholders, and technical experts in the MNH field.

## Methods

### Program context and description

The CBMNC Program was implemented in Aweil East County, Northern Bahr El Ghazal State, South Sudan situated along the border with Sudan. The program selected four *bomas* (the smallest administrative unit in South Sudan) for implementation, aligned with the ongoing implementation of the Boma Health Initiative (BHI), a national framework designed to strengthen community-level health service delivery in South Sudan [9]. This region is characterized by persistent food insecurity, recurrent disease outbreaks, conflict-related disruptions, and hosts significant populations of returnees and refugees from Sudan [10]. Access to health services remains extremely limited due to poor infrastructure, cultural norms and perception of low risk, and inadequately trained health personnel, making community-based MNH interventions both critical and challenging [11,12].

The Boma Health Workers (BHWs), the national community health worker cadre, employed by the BHI mainly delivered Integrated Community Case Management (iCCM) for Childhood Illness services prior to the introduction of CBMNC. The CBMNC program, which aligns with the BHI’s safe motherhood module, introduced key maternal and newborn health commodities alongside evidence-based practices. During the programme period, changes in the national BHW programme linked to a donor transition reduced the number of BHWs assigned per boma from 20 to 8, impacting the number of BHWs engaged in CBMNC. A summary of the CBMNC program’s structure and components is provided in Table 1.

**Table 1:**
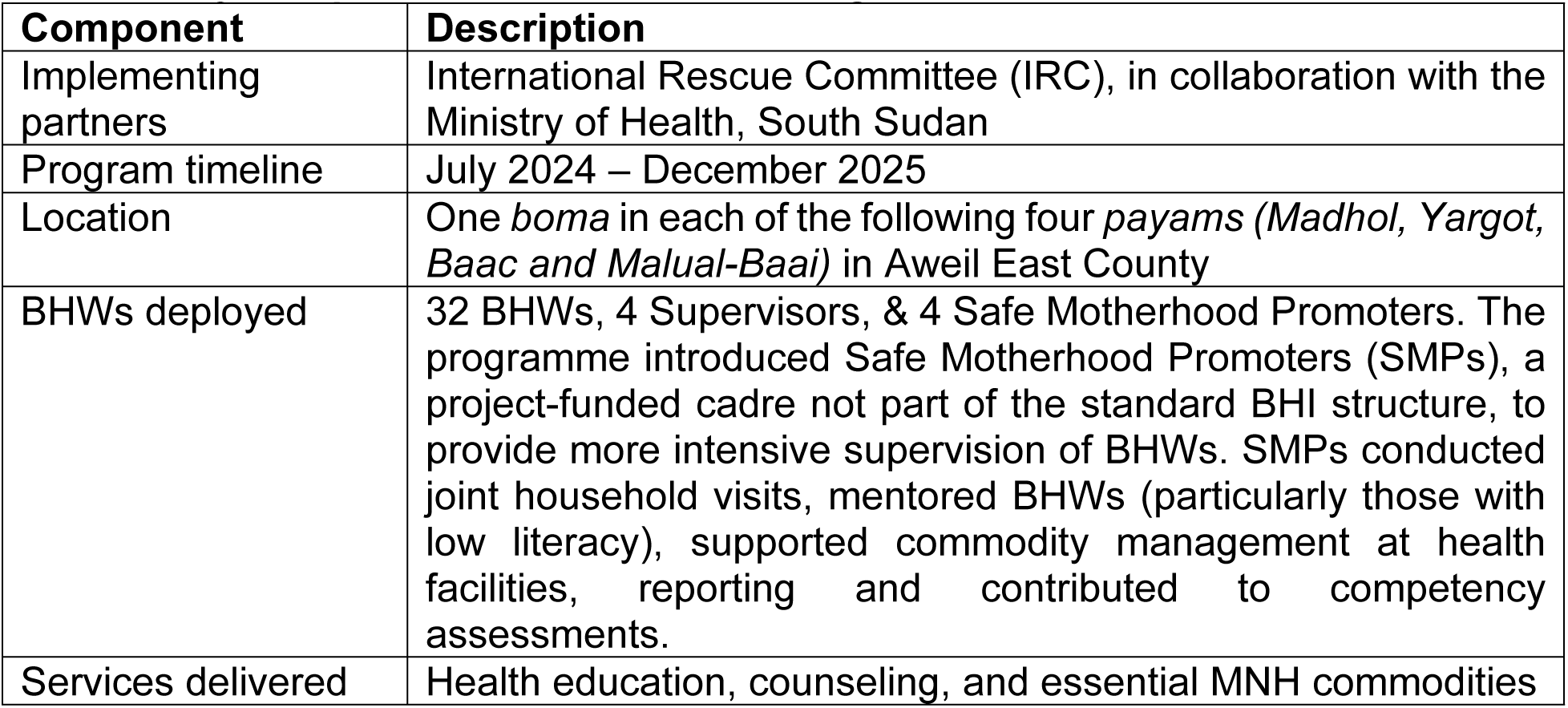
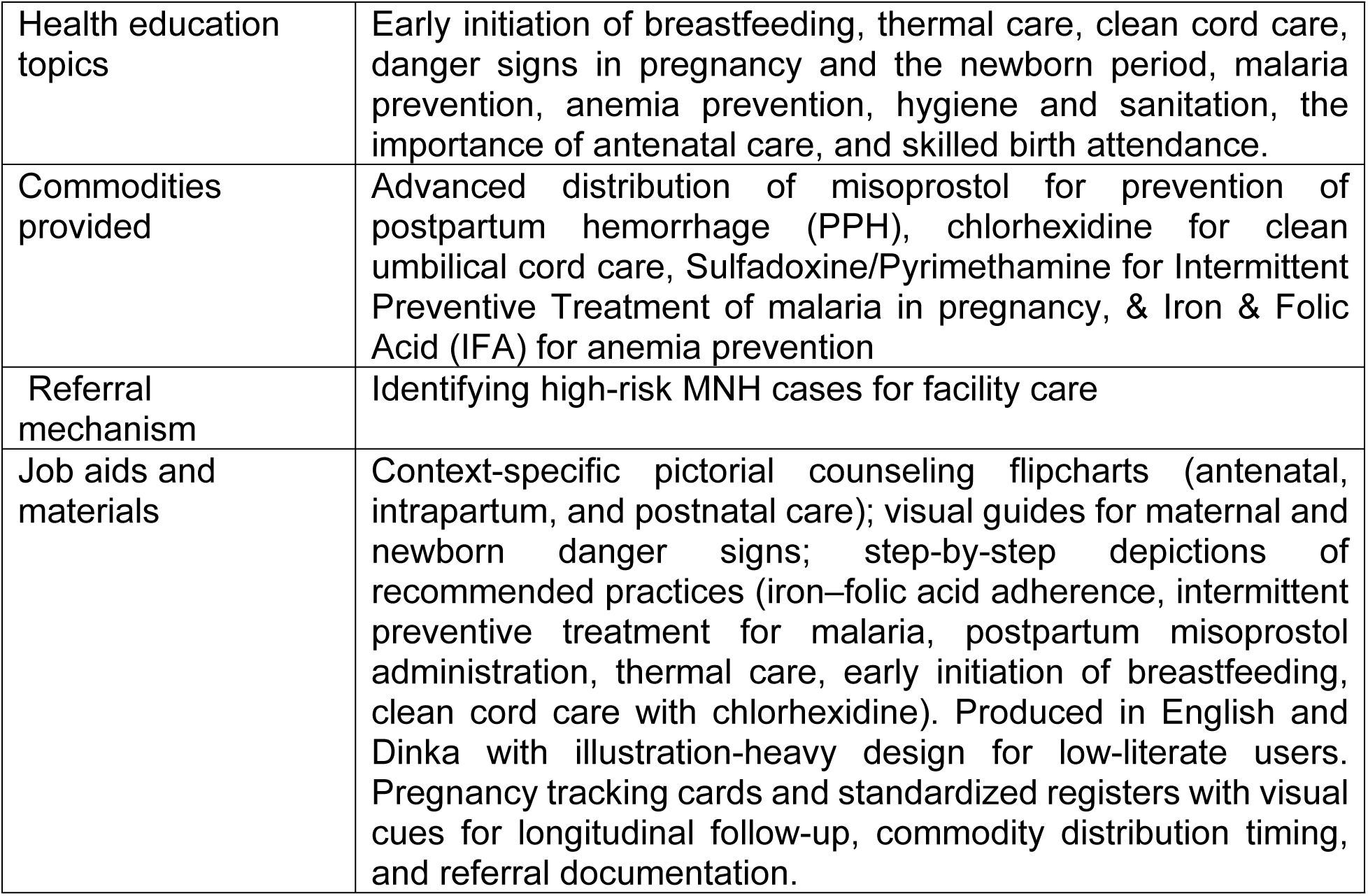
Key components of the CBMNC Program.

### Study design

This qualitative descriptive study explored the acceptability of the CBMNC program among diverse stakeholders in South Sudan. The study was grounded in the Theoretical Framework of Acceptability (TFA), which informed both the development of the study tools and the analytical approach [8]. Table 2 outlines the key constructs and their descriptions used in this study.

**Table 2:** Constructs described in the Theoretical Framework of Acceptability (Sekhon et al., 2017).

| Construct | Description |
| --- | --- |
| Intervention coherence | The degree to which participants understand the intervention and how it works |
| Affective attitude | How individuals feel about the intervention |
| Ethicality | The extent to which the intervention aligns with an individual's value system |
| Self-efficacy | Participants' confidence in their ability to perform the behaviors required by the intervention |
| Burden | The perceived effort required to participate |
| Opportunity costs | The benefits, profits, or values that must be forgone to engage in the intervention |
| Perceived effectiveness | The extent to which the intervention is perceived as likely to achieve its purpose |

### Study participants

The study included both service users and implementers to comprehensively explore program acceptability. Participants were purposively selected to ensure diversity in age, parity, disability status, geographic access, and level of program engagement. Key informants were identified through stakeholder mapping based on their roles in maternal and newborn health. FGDs explored shared perceptions and collective experiences, IDIs captured individual narratives and personal experiences, and KIIs provided systemic perspectives on program design and scalability. In total, 185 individuals participated (Table 3).

**Table 3:** Study participants by method, group, and sample size.

| Method | Participant group | Sessions | n |
| --- | --- | --- | --- |
| **FGD** | Women who received CBMNC services (clients) | 4 | 38 |
|  | Male partners | 4 | 34 |
|  | Mothers and mothers-in-law | 4 | 41 |
|  | Boma and Hospital Health Committee members | 4 | 40 |
|  | Supervisors and Safe Motherhood Promoters | 1 | 8 |
|  | **Subtotal** | **17** | **161** |
| **IDI** | Program recipients |  | 6 |
|  | BHWs |  | 8 |
|  | **Subtotal** |  | **14** |
| **KII** | State and national stakeholders |  | 10 |
| **Total** |  |  | **185** |

### Data Collection

Data were collected between May and July 2025. Interview and FGD guides were developed based on the TFA, tailored to specific participant groups, and explored participants’ understanding of the CBMNC program, perceived challenges, community perceptions, and contextual barriers. All data collection was conducted by trained qualitative researchers fluent in English and Dinka, who completed a five-day training program prior to fieldwork. Community-level interviews and FGDs were conducted in Dinka, while KIIs were conducted in English or Dinka based on participant preference.

Interviews were conducted in private locations selected in consultation with participants, while FGDs were held in neutral, accessible community spaces. Interviews lasted approximately one hour and FGDs approximately two hours. All sessions were audio-recorded with participant consent and supplemented by field notes. Saturation was assessed through concurrent thematic coding during data collection, with the research team pausing at regular intervals to review whether new themes were emerging. Data collection was concluded when successive interviews yielded no new themes across participant groups. Transcripts were produced verbatim in English by two bilingual transcribers and cross-checked against original audio recordings for accuracy.

### Data analysis

The study utilized a framework analysis approach, combining deductive coding, informed by the TFA, with inductive coding to identify emergent themes beyond the predefined domains. An initial codebook, structured around TFA constructs, was iteratively refined as new insights emerged during the analysis. Six researchers (LAL, TM, AD, ALA, GK, & JMD) conducted coding in Dedoose [13]. To establish coding consistency, all six researchers independently coded the same transcripts, then convened to compare code application, discuss discrepancies, and refine code definitions until consensus was reached. Although the TFA provided the primary interpretive lens, key themes were generated inductively to capture nuanced perspectives of service users and implementers regarding program acceptability.

### Ethical approval

This study received ethical clearance from the International Rescue Committee Institutional Review Board and the South Sudan Ministry of Health Research Ethical Review Board. Written informed consent was obtained from literate participants. For participants unable to read or write, informed consent was obtained verbally in accordance with approval granted by the relevant ethics committee. This approach was justified by low literacy levels in the study population and is consistent with international ethical standards for research conducted in such contexts. A standardized script was used to ensure that all participants received consistent information regarding the study objectives, procedures, potential risks and benefits, their right to withdraw voluntarily, and the confidentiality of their data. Participants were given the opportunity to ask questions before providing consent. Verbal consent was documented on a structured form capturing the date, participant name, and confirmation of agreement, which was then signed by the facilitator.

Adolescents aged 15–17 years were eligible for inclusion in the study. In the context of South Sudan and its Ministry of Health research ethics review board, those between the ages of 15-17 are eligible to provide informed consent without parental consent. Under South Sudan’s National Family Planning Policy (2013) and Reproductive Health Strategic Plan (2019–2023), adolescents are recognized as having the evolving capacity to make informed decisions about their reproductive health. Both the South Sudan research ethics board as well as the International Rescue Committee IRB (which abides by U.S. federal regulations) approved having children between ages 15-17 provide informed consent, without the requirement for parental or guardian permission, based on the content of the study and on the privacy protections we offered. Please refer to other publications that have also treated children between ages 15-17 as being able to provide informed consent / considered them emancipated minors [14,15].

### Reflexivity

This study was conducted by a team embedded within the implementing organization, which presents an inherent tension between organizational familiarity and research independence. The team was attentive to the ways organizational affiliation may have shaped data collection and analysis. Data collection was led by four South Sudanese researchers (two women and two men) who had no direct role in program implementation, but some participants may have had prior contact with personnel from the same organization. Researchers had no prior relationship with respondents except for one researcher who had existing professional relationships with some key informants. In those KIIs, this familiarity may have facilitated more candid responses rather than socially desirable ones. To address risks of social desirability bias and concerns about any consequence of respondent participation, researchers explicitly communicated during informed consent that the study sought genuine experiences and that responses would have no bearing on participants’ access to services or employment. Data analysis and triangulation were carried out by one of the researchers who collected the data and two researchers based out of Nairobi, neither of whom was involved in day-to-day program implementation, providing a degree of analytical distance. Findings were triangulated across respondent types and against available program context and documentation, to identify where accounts converged or diverged. Analysis actively sought disconfirming evidence to challenge assumptions arising from organizational familiarity. Where responses appeared to be shaped by social desirability, such as overly positive accounts of program experiences or overly negative in relation to financial or commodity provisions by the organization, they were retained in the analysis but interpreted with caution, ensuring such data points informed interpretation treating them as representative of broader participant experience.

## Results

The findings are organized according to the seven TFA constructs (Table 2).

### Intervention coherence

Most stakeholders understood what services were offered, how to use them, and why they mattered, though comprehension varied by literacy, prior experience, and proximity to service delivery. Across all groups, CBMNC was understood as bridging geographic gaps rather than replacing facility care. As one BHW explained: *“I emphasize that going to the health facility is paramount for every pregnant mother”* (IDI-BHW). Male partners similarly recognized that serious complications required facility care.

Some distal health system stakeholders worried communities might see this as the complete care package, potentially discouraging facility visits*: “Some community members may think that this is the whole package they should receive”* (KII-National). However, local facility staff did not observe any negative reductions in care-seeking behavior.

Understanding was notably stronger for medications than for health education messages. Participants could describe each medication’s purpose; for example, *iron-folic acid “increases blood in the body,” misoprostol “stops bleeding,” and chlorhexidine “quickly heals the cord”* (FGD-Clients). Iron-folic acid and antimalarials were familiar due to prior community exposure, while chlorhexidine and misoprostol required more explanation because they were *“new to us”* (FGD-Clients). Committee members could describe timing instructions with precision. In contrast, health education topics such as breastfeeding, thermal care, and danger sign recognition were rarely discussed.

Misoprostol raised particular concerns. Some younger and first-time mothers associated it with abortion rather than postpartum bleeding prevention. BHWs addressed these fears through repeated clarification, emphasizing use *“only after delivery”,* and described precise safety protocols: *“I instruct them to take it only after they are sure the baby is fully delivered and no twin is left inside the womb”* (IDI-BHW).

Understanding was built through multiple reinforcing pathways that compensated for low literacy. Clients identified medications by color, packaging, and timing rather than written instructions: *“We lay them out on the floor, and try to recall the explanations, matching each drug to what was said”* (IDI-Client). Repeated BHW visits were critical, with understanding deepening across interactions: *“The second visit is connected to the first. So, she does not take time if she sees that you have followed her instructions well” (*IDI-Client).

Social learning also played a key role. Male partners learned by observing their wives’ experiences, and mothers-in-law developed enough practical knowledge through repeated observation to support medication use during home deliveries*: “I removed the three tablets and gave them to her… and I removed the other medication and applied to the umbilical cord”* (FGD-M/MIL). At the provider level, pictorial job aids compensated for literacy limitations among BHWs, helping guide discussions with clients who could not read.

### Affective attitude

Participants from all stakeholder groups felt positively about the program, expressing feelings of relief, reassurance, gratitude, and emotional security, but also expressed concerns about sustainability. Providers also experienced frustration due to insufficient support.

Clients described feeling emotionally supported by BHW presence during pregnancy and the postnatal period. Regular home visits, counselling, and medication availability reduced anxiety related to complications, distance to facilities, and uncertainty around childbirth, particularly for women who had previously experienced pregnancy-related complications. Emotional trust was central to these responses: women emphasized feeling comfortable discussing sensitive health matters without fear or shame, and the perception of BHWs as “like a brother or a sister” facilitated open communication and strengthened intervention uptake.

Male partners described CBMNC as alleviating longstanding anxieties about pregnancy-related complications and the financial burden of emergency facility care, with personal testimonies of wives’ safe deliveries generating strong positive affect*: “My wife, for instance, who was enrolled and kept on taking them [medicine], delivered well and healthy… Her delivery was never easy in the past, and so I believed it was the medications that made her healthy, resulting in a safe delivery”* (FGD-Male Partners). Mothers and mothers-in-law expressed similar relief framed through intergenerational comparison, positioning CBMNC as addressing traumas from their own childbearing years*: “If these drugs had been available in the past, many of our children would have survived… Why did these good services come only now, when we are old?”* (FGD-M/MIL).

BHWs expressed satisfaction in perceiving improved maternal and newborn outcomes and receiving community recognition. However, positive effects were severely constrained by frustration over inadequate compensation, as BHWs received 50 USD per month for BHI services with an additional 15 USD for CBMNC, and by their inability to meet client expectations beyond their mandate. One noted: *“What we get from this work is not enough to provide food to the family… we accept to work because the community that is being supported is ours and the children are our children, too, despite the challenges*“ (IDI-BHW). Despite these challenges, BHWs continued their work, motivated by communal responsibility and faith: “Even if I don’t get anything, God will reward me elsewhere” (IDI-BHW). BHC/HHC members similarly felt undervalued as unpaid volunteers, yet positioned themselves as program guardians: *“We are the batteries of the community and if we hide the reality that is on the ground in the community, then it is like a torch that has no batteries” (FGD-BHC/HHC)*.

Health system stakeholders were broadly supportive, noting that CBMNC reduced facility workload by moving routine commodity distribution to the community level. One stated: *“Many women come only to collect iron supplements, Fansidar, or deworming tablets. If these could be accessed at the community level, it would ease our workload significantly”* (KII-State). Misoprostol drew particular concern from distal health system stakeholders at state and national levels regarding safe distribution by non-clinical providers, though no direct observation or knowledge of misuse was raised by any respondent and community-level actors expressed confidence in the safety protocols in place.

### Ethicality

CBMNC was uniformly regarded as morally suitable and consistent with community values. All stakeholder groups framed CBMNC as *“the right thing,”* rooted in recognition that remote location, poverty, and absent health infrastructure created conditions where women died from treatable complications. One male partner stated: *“What the program has done is incredible because where we live is a village with no access to medical services… CBMNC is a health intervention brought to address this gap” (FGD-Male Partners).* BHWs described their work as a humanitarian obligation, while clients and male partners compared CBMNC favourably with costly facility-based or traditional birth attendant services.

Initial skepticism from some elders about home-based medication distribution required negotiation: *“They questioned why medications were being distributed and taken at home instead of at the health facility. They believed that medicines given outside the health center might not be effective*“ (IDI-Client). Women convinced elders through their own positive experiences. Mothers and mothers-in-law framed the replacement of harmful traditional practices such as applying cow dung or lizard droppings to the umbilical cord as moral progress: *“Harmful traditional practices have been replaced… Today, health workers give mothers proper medicines”* (FGD-M/MIL).

Women expressed comfort discussing intimate health matters with female BHWs, while cultural norms constrained male BHWs’ scope for example, they could not demonstrate breastfeeding positioning techniques. BHWs’ respectful conduct and confidentiality established them as trustworthy: women’s belief that *“a doctor cannot bring medication that kills a patient”* (FGD-Client) extended to accepting unfamiliar medications. Community selection of BHWs reinforced this trust, creating reciprocal obligations: *“we accept to work because the community that is being supported is ours and the women and the children are our children, too, despite the challenges.” (IDI*-BHW)

Despite this trust, concerns about fairness arose. Multiple groups questioned why services excluded other populations. Male partners asked: *“Are men not also at risk of anemia?… So why are there no drugs for men in such conditions?”* (FGD-Male Partners). BHC/HHC members expressed discomfort when neighboring villages were excluded: *“People complain a lot that the services are based here and only meant for [specific community] children”* (FGD-BHC/HHC).

### Self-efficacy

Participants generally showed high self-efficacy, though it varied by task complexity, literacy, and support systems.

Clients expressed high confidence in adopting recommended practices such as breastfeeding, thermal care, and clean cord care, particularly when instructions were clear, repeated, and delivered in local language. Self-efficacy for medications was more conditional, depending on instruction clarity and BHW reassurance; non-literate women relied on visual differentiation and memory for correct medication identification. Repeated BHW visits progressively strengthened confidence, and clients described returning to BHWs for clarification without shame. Male partners and mothers-in-law positioned themselves as secondary monitors supporting medication adherence*: “Our role during the pregnancy of our daughters is to ensure that they take the medicines that the health personnel has given”* (FGD-M/MIL). Those with traditional birth attendant experience expressed confidence administering commodities during home deliveries, while others preferred BHW presence.

Among providers, BHWs expressed high confidence in perceived straightforward tasks like Fansidar distribution but lower confidence for socially sensitive medications. Misoprostol generated particular concern around accountability, as the abortion associations described under Intervention coherence carried legal implications in South Sudan: *“I give this misoprostol with fear because if it happens that the instructions don’t get followed and any implications happens, I will be blamed”* (IDI-BHW). BHWs developed safety strategies such as giving instructions when the spouse was present. Literacy limitations created variable self-efficacy for administrative tasks; despite adapted job aids, illiterate BHWs relied on memory and supervisory support: *“I don’t know how to read or write, so I rely on my memory while I’m in the field”* (IDI-BHW).

### Burden

Clients experienced minimal effort needed to participate in the program. Home-based service delivery eliminated the physical demands of travelling to distant facilities during pregnancy. One explained: “*Pregnancy often comes with fatigue, and even walking short distances can be exhausting for a woman. Having the medications delivered directly to her home makes a significant difference*” (FGD-Client). Men noted that home visits also removed the risks associated with travel: *“Receiving the medications from far has the risks associated, especially if it involves the use of the motorbike, which has a high risk of accidents*” (FGD-Male Partners). Brief visit duration (typically 30 minutes to one hour, shorter for follow-ups) further minimized effort, and BHWs’ practice of visiting during morning hours reduced disruption to daily routines. However, hospitality expectations created modest social demands for some extremely poor households: *“When they [BHWs] come as visitors, you, as the owner of the household, would feel welcoming and give them tea”* (IDI-Client).

BHWs however experienced substantial physical, temporal, and economic burdens that they perceived as not proportional to the financial compensation for their work. Walking vast distances to scattered households dominated the narratives. One stated: *“The main challenge is the means of mobility as households are scattered, and walking is tedious… I end up delivering these medications until evening and return home”* (IDI-BHW).

Each BHW was responsible for covering 40–80 households spread across multiple villages, an increased ratio due to change in national policy that assigned 20 BHWs per boma to 8 BHWs per boma during the course of the project (the policy has since reverted back to 20 BHWs per boma). Another described scale: *“I have 7 villages under my Boma where I deliver medications to the mothers… A single household takes about 15-30 minutes [to conduct the follow-up activities] before moving to the next household”* (IDI-BHW). The work consumed entire days: “…*on my working day, I don’t do anything else in my house because the villages are vast and the households are scattered”* (IDI-BHW). Weather and terrain compounded seasonal physical demands without protective equipment. BHWs described working through rain, mud, and thorn: “*Sometimes, when it rains while I’m in the field, I have to take shelter in someone’s home until the rain stops, and then I continue with my work*” (IDI-BHW). BHC/HHC members also shared physical demands, with multiple participants describing multi-hour walks to attend meetings.

Client expectations for material goods beyond what is provided by the program created a psychological burden. BHWs described shame and stress from unmet requests: *“It is a challenge because I am taken like a doctor who has money by the mothers as some of them ask for money for soaps and I feel so ashamed when I reply that there is nothing”* (IDI-BHW).

Health system stakeholders recognized the intensity of provider effort as a sustainability concern, noting frustration with administrative complexity and coverage demands: “*They [BHWs] say, ‘we are overloaded with more paperwork*’” (KII-State). For health facilities, stakeholders perceived CBMNC as burden-reducing by managing cases at the community level, though supervision burden on county and state staff was acknowledged as substantial.

### Opportunity costs

Opportunity costs, defined as the benefits, profits, or values forgone in order to participate, reflected the pattern of burden: they were minimal for service recipients but considerable for providers.

For service recipients, opportunity costs were minimal. Activities paused to receive CBMNC services like farming, food preparation, water collection were readily resumable, and time spent was framed as a worthwhile investment: “*If I am found doing any work and a health worker arrives, I immediately stop what I’m doing and ask my mother to take over*” (IDI-Client). Mothers-in-law also absorbed opportunity costs by assuming household tasks to enable daughters’ service access.

BHWs however experienced substantial, multidimensional opportunity costs encompassing foregone agricultural productivity, household income, and family care time. Farming sacrifice during critical seasons was prominent, with supervisors observing: “*These women now among them don’t cultivate during the farming season*” (FGD-BHC/HHC). BHWs described prioritizing livelihood needs over programme duties when trade-offs became acute: “*During the rainy season, I prioritize planting over drug distribution… I might not be able to complete what I had planned [for BHW work]*” (IDI-BHW). Household management time losses created cascading costs for some families, particularly BHWs with young children: “*I’m currently carrying a newborn, which makes the work more difficult”* (IDI-BHW).

Compensation was insufficient to offset these foregone values. BHWs received $15 USD monthly, which was rapidly depleted: “*We are paid only 80,000 South Sudanese Pounds (SSP) [$15], and that money runs out in just two or three days… We constantly move around, trying to balance this volunteer work with other small jobs*” (IDI-BHW). Supervisors received an additional $25 monthly top-up for CBMNC duties but still bore out-of-pocket expenses: “*During one of our field visits, my colleague and I had to pay SSP 40 per trip to cross the river using a boat… If BHWs are expected to cover these transportation costs from their modest remuneration, they are left with almost nothing*” (FGD-Supervisor).

BHC/HHC members described leaving livelihood activities, market sales, and farming to support CBMNC activities without compensation. Their extensive unpaid work conducting monitoring, mobilization, and emergency response represented a significant sacrifice, particularly during farming seasons.

Health system stakeholders recognized inadequate compensation relative to opportunity costs as a sustainability threat. Some BHWs managed by working with multiple organizations simultaneously.

### Perceived effectiveness

All stakeholder groups perceived CBMNC as effective, drawing on direct experience and comparisons with outcomes before the programme.

Commodity-based interventions produced the strongest perceptions of effectiveness. Misoprostol was most frequently cited, with clients, partners, providers, and facility staff all attributing reductions in postpartum bleeding to the drug. One client recounted: *“I am a mother of 6 children, and three of the deliveries had complications when placenta delayed… the birth of the 6th baby was the best with no complications because of the CBMNC medications”* (FGD-Client). Supervisors perceived that PPH-related deaths had not occurred in coverage areas, and facility staff corroborated that deaths “*still happen*” in areas not covered by the CBMNC program while covered areas experienced none. Chlorhexidine was associated with faster cord healing, iron–folic acid with reduced anaemia, and Fansidar with fewer malaria episodes during pregnancy.

Behavioural counselling was mentioned less frequently and in less specific terms. Supervisors observed changes in newborn care practices such as delayed bathing and skin-to-skin contact, but clients rarely cited counselling as evidence of programme impact. BHWs described limited success changing behaviours around facility delivery as well as referral completion.

These perceptions extended beyond individual experience to community-wide observations of reduced maternal mortality and morbidity and drove strong advocacy for programme expansion: *“We have accepted the medications as community of [village name] but please increase the number of the BHWs”* (FGD-BHC/HHC).

Beyond health outcomes, participants described improvements in daily life. Women described reduced anxiety, increased mobility, and restored capacity for daily activities. BHWs reported clients noting: *“This time I didn’t bleed like before. I can cook and eat on my own”* (IDI-BHW). Women repeatedly expressed concern that health gains would be reversed if services stopped: *“the suffering is going to come back only when this medication is stopped”* (FGD-Client).

Finally, health system stakeholders believed in the effectiveness of CBMNC interventions but expressed variable confidence in BHW capacity, contingent on task complexity. Confidence was high for straightforward tasks like IFA and Fansidar distribution and for counselling on practices like breastfeeding. Consistent with the concerns noted under Intervention coherence and Self-efficacy, misoprostol generated divided perceptions: state-level stakeholders expressed conditional confidence given the absence of reported incidents, while national-level stakeholders maintained doubt about BHWs’ ability to deliver it safely. Stakeholders emphasised the need for systematic evaluation data before informing scale-up decisions.

## Discussion

Our findings contribute to the limited evidence on community-based maternal and newborn health interventions in conflict-affected settings, where a disproportionate share of global maternal and newborn deaths occur [16]. Our study applied the TFA systematically across seven constructs of acceptability, revealing not only that CBMNC was accepted but how and why from the lens of multiple stakeholder groups.

### Trust as the foundation of acceptability

Trust and relational bonds with BHWs were central to acceptability. Women described BHWs as *“like a brother or a sister”* and attributed their children’s survival directly to individual providers. This aligns with established evidence on the role of CHWs as intermediaries bridging health systems and communities, and on trust as a driver of health-seeking behaviour in sub-Saharan Africa and fragile contexts specifically [3,17–19]. Research from post-Ebola Guinea found that provider competence, empathy, and confidentiality were core to rebuilding trust and restoring service use [20], all of which BHWs appeared to demonstrate.

Community selection of BHWs was foundational to this trust, creating a sense of mutual obligation between providers and the communities they serve. This aligns with evidence that community selection and engagement of local leaders are crucial for acceptance and use of CHW services in humanitarian settings [21]. Programme designs that bypass community selection in favour of efficiency risk undermining this relational foundation.

Male partner engagement also supported acceptability, with partners who understood interventions providing permission, support, and encouragement for medication adherence. This is consistent with evidence that male engagement improves uptake in settings where men hold decision-making authority over women’s healthcare [1,2]. However, partner engagement was primarily framed around commodities rather than behavioral counselling, reinforcing the gap in visibility discussed below.

Beyond individual relationships, CBMNC addressed emotional dimensions of care-seeking that are often overlooked. Women described profound relief that services came to them, removing the need for costly and sometimes dangerous journeys to distant facilities. This sense of security may be as important to sustained engagement as the interventions themselves.

### Achieving understanding in low-literate populations

Clients, male partners, and mothers-in-law demonstrated sufficient understanding for safe medication use despite widespread low literacy. This was achieved through multiple reinforcing channels: pictorial job aids, repeated BHW visits, and learning from peers. These findings have practical implications for building multiple pathways for comprehension.

A related question not explored in this programme is whether more biomedical explanations of interventions can be made accessible to low-literacy populations. Research from Kenya and South Africa suggests that where biomedical explanations are culturally grounded, they can coexist with and even replace traditional frameworks, strengthening both comprehension and demand [22,23]. Investing in accessible technical explanations, rather than assuming they are beyond the population’s reach, may further strengthen understanding.

Notably, health system stakeholders expressed concerns about whether low-literate providers could safely tell medications apart. Yet reports from clients, BHWs, and facility staff did not support these concerns. While these are perceived rather than objectively verified outcomes, their consistency across independent stakeholder groups suggests that policymaker hesitancy may not be warranted when appropriate training, supervision, and support tools are in place. Systematic documentation of safety outcomes is needed to confirm this and inform scale-up decisions.

### The salience gap between commodities and behavioral counselling

Understanding and perceived effectiveness were notably stronger for commodity-based interventions than for behavioral counselling. This is consistent with evidence that behavioral interventions are harder to perceive as impactful when benefits are gradual or not immediately visible [24,25]. Importantly, lower visibility in participant accounts does not necessarily mean lower effect. Gradual, diffuse benefits are inherently harder to recall and describe than the immediate, visible outcomes of medications.

Evidence suggests that the perceived value of behavioral interventions increases when reinforced through social proof rather than individual counselling alone, when delivered by trusted community-selected providers and when embedded in repeated, concrete interactions [26,27]. Some of these mechanisms were present in CBMNC, yet their impact was stronger for commodity uptake. In this programme context, the gap carries practical risks: if women cannot readily articulate the value of counselling, adherence to behavioral components may erode over time, and community demand for these services may be weaker when programmes seek to scale or sustain funding. Future programme design should consider how to extend these reinforcing mechanisms to counselling components, including deliberate strategies to make gradual behavioral gains visible to participants, such as structured milestone tracking or community-level outcome sharing. Few studies have compared how clients perceive the value of commodity-based versus behavioral interventions within the same programme in humanitarian settings, and this is an important area for further research.

### Provider burden and the acceptability paradox

A critical tension emerged: high community acceptability rested substantially on providers absorbing physical, time, and economic costs that were not adequately compensated. The effort required to build trust through repeated home visits, reassurance, and maintaining confidentiality is a form of provider burden rarely captured in workload assessments, yet this investment appeared to drive the acceptability to communities. Literature suggests minimally compensated models produce high turnover and quality risks [28,29], and several respondents spoke to BHWs working for multiple programs simultaneously for compensation. The World Health Organization Guidelines on Health Policy and System Support to Optimize Community Health Worker Programs recommends remunerating CHWs with a financial package commensurate with their job demands, hours, training, and scope of work [30]. The South Sudan’s Boma Health Initiative, the community health policy, reflects high ambitions in relation to the number and types of service delivery modules a BHW could deliver. The policy does not offer guidance on how compensation is to be adjusted as mandates differ by program; as more opportunities arise like the CBMNC program to expand the life-saving potential for a community health cadre, more standardized guidance should be provided on adjustments to compensation or a cap on workload to match the compensation available.

### Sustainability

Participants’ concern about programme discontinuation and stakeholders’ concerns about dependency on external funding underscore the need for sustainability planning from the outset. A review of CHW programme scale-up identified three prerequisites: effective programme management, community fit, and integration with the broader health system [29]. CBMNC met the second criterion strongly and the first partially, but the third, systems integration, remains the most significant gap. While embedding CBMNC within the BHI provides a pathway, sustainable scale-up requires government commitment to CHW compensation, supply chains, and supervision that extends beyond donor funding cycles. Sustainability also requires closing gaps between policymaker and community perceptions of acceptability. Misoprostol illustrates this clearly: respondents most proximate to the service raised minimal concerns about community-level delivery, while those most distal expressed the greatest hesitancy. A political economy analysis conducted on prioritization of MNH in South Sudan reflects on inadequate inclusivity of women and girls in policymaking, leading to “policies that inadequately capture the perspectives of the affected population, both as decision makers and as recipients of services [31]. When policies dictate access for women and newborns to life-saving services, there is a critical imperative to ensure these perception gaps are closed with both the lived experience of the provider and client, as well as concrete evidence.

### Strengths and limitations

The comprehensive application of the TFA across multiple stakeholder groups provides a detailed understanding of acceptability that is often missing from evaluations. Several limitations warrant consideration. The study was conducted in a single geographic area, limiting generalizability. Data were collected midway through the implementation period (July 2024–December 2025), and perceptions of effectiveness and provider burden may evolve as the programme matures. Social desirability bias may have influenced responses, particularly given that data were collected by staff from the implementing organization, though the research team took precautions in data collection, analysis, and interpretation to minimize its impact. Selection of participants through programme records may have excluded women who declined services, potentially biasing findings toward more positive perspectives.

## Conclusion

CBMNC was well accepted when delivered through trusted, community-selected health workers using contextually appropriate strategies. Ways to minimize the gaps in visibility to both client and provider between commodity-based and counseling-based interventions need to be deliberately designed for to further improve the health outcomes resulting from such programs. There also needs to be more attention paid to provider welfare, including adequate compensation, functional equipment, and manageable workloads, as a prerequisite for scale-up. For programmes like CBMNC to move beyond donor-funded pilots, governments and implementing partners must address the policy barriers that restrict community-level delivery of life-saving commodities such as misoprostol and commit domestic financing to CHW compensation and supply systems.

## Data Availability

Given more than minimal potential of identifiability for the KII transcripts conducted with key stakeholders at the state and national levels, we will have the transcripts available for all data in time for publication, excluding KIIs

## Acknowledgements

The authors wish to acknowledge Manga Benard Manga and Fred Innocent Ali of the International Rescue Committee South Sudan for their invaluable support in the coordination and implementation of field activities that made this study possible

## Supporting information

**S1 File.** S1_Cover letter.pdf

**S2 File.** S2_Human Participants Research Checklist.pdf

**S3 File.** S3_MoH Ethics Clearance2024.pdf

**S4 File.** S4_MoH Ethics Clearance2025.pdf

**S5 File.** S5_IRC Ethics Clearance2024.pdf

**S6 File.** S6_IRC Ethics Clearance2025.pdf

